# THE PREVALENCE AND FACTORS ASSOCIATED WITH INTIMATE PARTNER VIOLENCE AMONG PREGNANT WOMEN IN TANZANIA: A CROSS-SECTIONAL ANALYSIS OF THE 2022 TANZANIA DEMOGRAPHIC AND HEALTH SURVEY AND MALARIA INDICATOR SURVEY (2022-TDHS-MIS)

**DOI:** 10.64898/2026.03.13.26348362

**Authors:** Cephlen Mathayo, Rose Mpembeni, Jasmine Chilembu, Agnes Tesha, Godson Ngowi, Rogath S. Kishimba, Habib Ismail, Salum Faru, Jonhas Masatu

## Abstract

**Background:** Intimate partner violence (IPV) during pregnancy is a critical public health and human rights issue that affect almost 30% of women and threatens maternal and fetal health among pregnant women. Despite the recognized burden of IPV in Tanzania, the prevalence and determinants among pregnant women need to be well identified using the national representative data.

**Objective:** This study aimed to determine the forms, prevalence, and factors associated with intimate partner violence among pregnant women in Tanzania using the 2022 Tanzania Demographic and Health Survey and Malaria Indicator Survey (TDHS–MIS) data.

**Methods:** A cross-sectional study analyzed secondary data from the 2022 Tanzania Demographic and Health Survey and Malaria Indicator Survey (TDHS–MIS) on intimate partner violence (IPV) among pregnant women aged 15 – 49 years. A total of 435 pregnant women who responded to the IPV module were included. Weighted descriptive statistics estimated prevalence and forms of IPV, while modified Poisson regression determined factors associated with IPV. Adjusted prevalence ratios (APRs) with 95% confidence intervals (CIs) were reported.

**Results:** The overall prevalence of IPV among pregnant women was 27.46% (95% CI: 22.94–32.50). Emotional violence was most common (25.26%), followed by sexual (11.04%) and physical (11.01%) violence. IPV prevalence was highest in Mara (60.3%), Songwe (50.1%), and Singida (39.0%) regions. Factors independently associated with IPV included partner alcohol use (APR = 2.55; 95% CI: 1.50–4.31), partner having other wives (APR = 1.75; 95% CI: 1.11–2.87), and union duration of 5–9 years (APR = 2.65; 95% CI: 1.14–6.18). Having a marriage certificate (APR = 0.51; 95% CI: 0.28–0.92) and one child (APR = 0.40; 95% CI: 0.17–0.95) were protective.

**Conclusions:** IPV affects more than one in four pregnant women in Tanzania, with emotional abuse being predominant. Partner alcohol use, polygamy, and longer union duration heighten IPV risk. Integrating IPV screening and counseling into antenatal care and implementing behavior change interventions for partners could reduce the burden of violence during pregnancy.

## Introduction

Intimate partner violence (IPV) remains a major public health and human rights concern worldwide. The World Health Organization estimates that about 30% of women globally have experienced physical or sexual IPV, and between 4% and 12% experience such violence during pregnancy. Despite global advocacy, the burden of IPV during pregnancy has shown little decline over the past two decades, particularly in low- and middle-income countries.

In sub-Saharan Africa, IPV during pregnancy remains widespread, with estimates ranging from 15% to 41%, and emotional abuse being the most prevalent form. Such violence increases the risk of miscarriage, preterm delivery, low birth weight, depression, and maternal death.

In Tanzania, IPV among women of reproductive age has remained persistently high 39% in 2010, 40% in 2015–16, and 39% in 2022 with limited disaggregated data for pregnant women. Facility-based studies have reported IPV prevalence among pregnant women between 25% and 30%, suggesting that pregnancy does not offer protection against abuse.

Understanding the magnitude and factors associated with IPV during pregnancy is critical for improving maternal health outcomes and achieving Sustainable Development Goals (SDG 3 and SDG 5). This study therefore examined the prevalence and determinants of IPV among pregnant women aged 15–49 years in Tanzania using data from the 2022 TDHS–MIS.

## Methods

### Study Design, Data Source and Access

This study employed a cross-sectional design using secondary data from the 2022 Tanzania Demographic and Health Survey and Malaria Indicator Survey (TDHS-MIS), a nationally representative cross-sectional survey implemented by the National Bureau of Statistics (NBS) and the Office of the Chief Government Statistician (OCGS) in collaboration with the Ministry of Health, with technical assistance from ICF through the DHS Program. The survey collected data between 24 February 2022 and 21 July 2022 using computer-assisted personal interviews.

The TDHS-MIS dataset used in this study was obtained from the DHS Program after formal approval. The dataset was accessed for research purposes on 14 August 2024.

The survey utilized a two-stage cluster sampling method to ensure national representativeness, selecting 629 enumeration areas (EAs) and 16,354 households across Tanzania Mainland and Zanzibar. A total of 15,254 women aged 15–49 years were interviewed, among whom 1,165 were pregnant. Only 435 pregnant women who responded to the IPV module were included in this analysis.

### Variables

**The dependent** variable was experience of IPV during the current pregnancy, defined as having experienced at least one form of physical, sexual, or emotional abuse by a partner.

### Independent variables

**Respondent characteristics:** age, education, marital status, duration in union, working status, number of children, place of residence, acceptance of wife beating.

**Partner characteristics:** age, education, occupation, alcohol use, and number of other wives.

**Household and social factors:** wealth index, area residence and region.

### Data extraction procedures

Secondary data was used in this study from the 2022 TDHS-MIS, a nationally representative survey that collected comprehensive demographic and health-related data, including data on intimate partner violence using the individual woman’s questionnaire. The permission to use the TDHS-MIS dataset was obtained from the DHS program secretariat with a letter dated on Aug 14, 2024, that allowed me to use the dataset from Tanzania.

From the 2022 TDHS-MIS dataset, a sample of 15,254 women were interviewed, of which 1167 women were pregnant at the time of the survey. Among the pregnant women, 435 were interviewed on the experience of violence by their husband or intimate partner during pregnancy. These 435 women formed the sample for the analysis of forms, prevalence, and factors associated with IPV among pregnant women in Tanzania as it is explained in figure 2 below.

**Figure 1:**
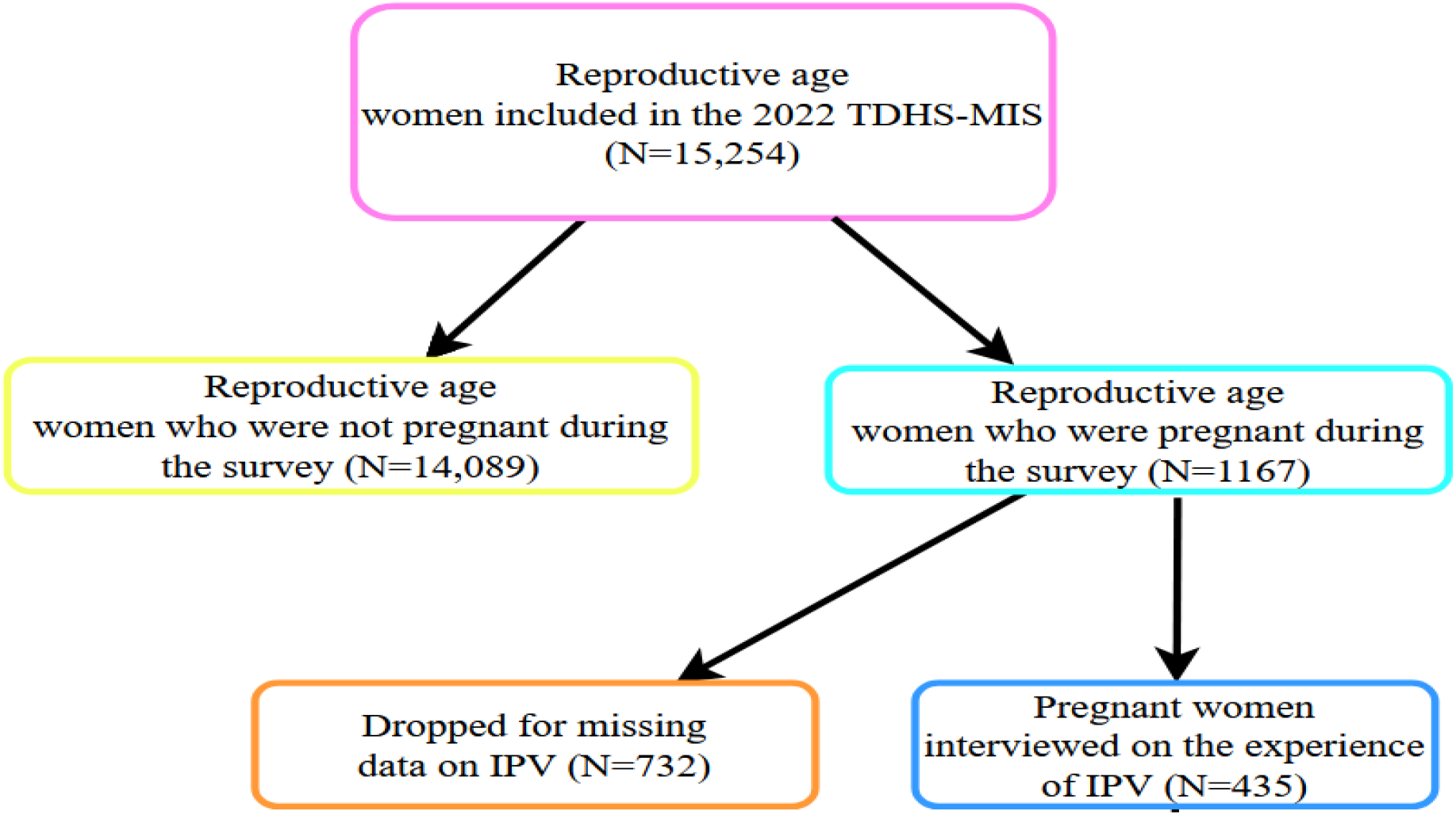
The data extraction procedure and sample size.

**Figure 2:**
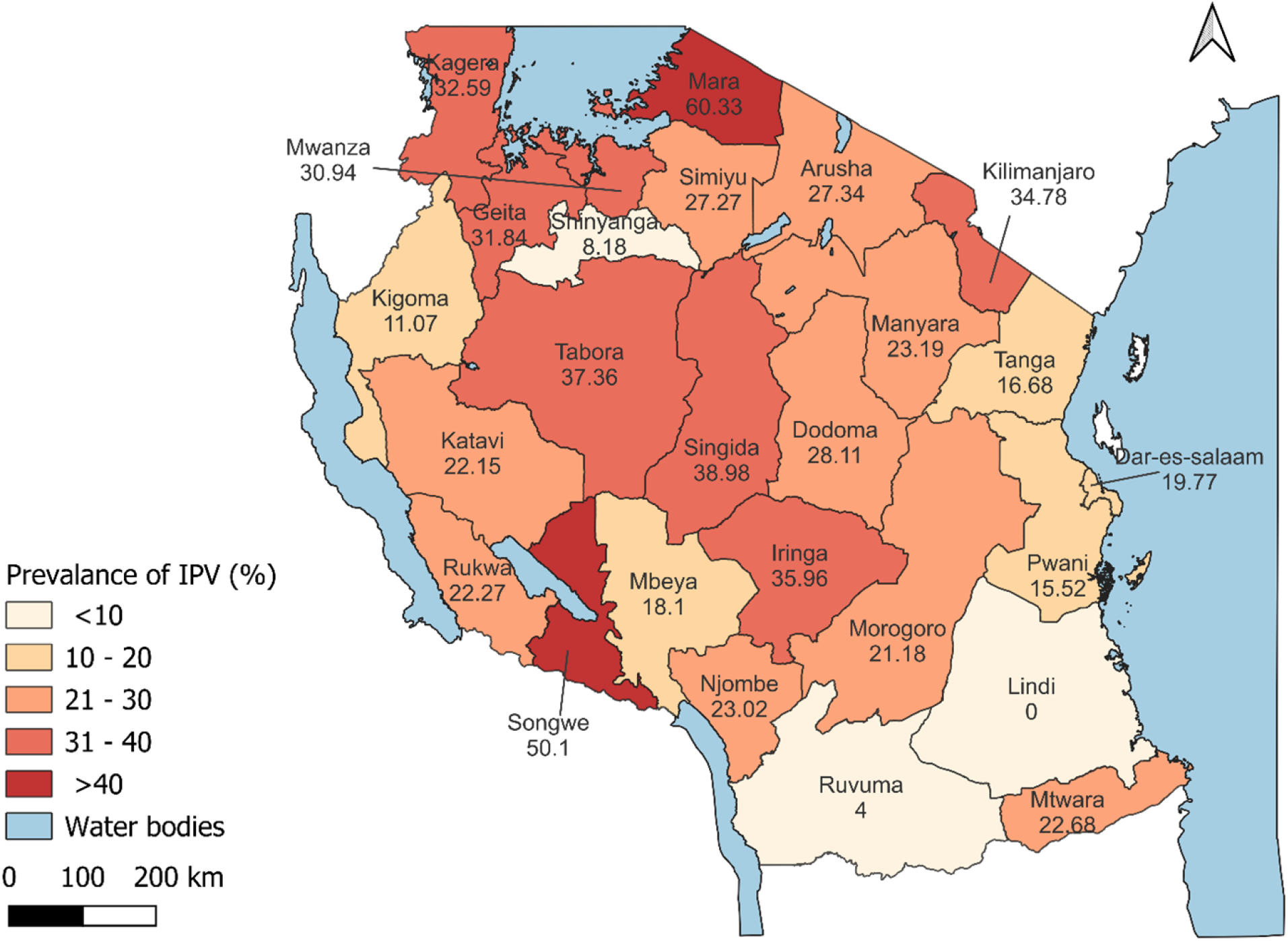
Map showing the prevalence of IPV among Tanzanian pregnant women aged 15 - 49 years

### Data Analysis

All analyses were conducted in STATA version 18, accounting for survey weights, clustering, and stratification using DHS commands (svyset). Descriptive analyses described socio-demographic characteristics and IPV prevalence. Modified Poisson regression was used to estimate crude and adjusted prevalence ratios (PRs and APRs) with 95% CIs. Variables with *p* < 0.20 in bivariable analysis or conceptual importance were entered into multivariable models. Statistical significance was set at *p* < 0.05.

### Ethical Considerations

Ethical approval was obtained from the MUHAS Research and Publications Committee (Ref. No. DA.282/298/01.C/2586). Permission to use TDHS–MIS data was granted by the DHS Program. The TDHS–MIS received ethical clearance from the National Institute for Medical Research (NIMR) and adhered to WHO guidelines for the ethical collection of IPV data.

### Confidentiality and access to identifiable information

The TDHS-MIS datasets are fully anonymized prior to release, and all personal identifiers such as names, exact addresses, and geographic coordinates are removed or displaced. Therefore, the authors did not have access to any information that could identify individual participants during or after data collection. The survey obtained informed consent from all respondents before participation.

## RESULTS

### Characteristics of Respondents

The median age of respondents was 27 years (IQR: 23–33). Over half (51.0%) had primary education, 72.4% resided in rural areas, and 69.4% were married. About 20.1% reported that their partners used alcohol, and 15.4% had partners with other wives. Most respondents (91.3%) disagreed that wife beating was acceptable.

**Table 1:** Socio-demographic characteristics of pregnant women who responded to the IPV questionnaire in Tanzania from the 2022 TDHS-MIS.

| Respondent Characteristics | Frequency | Percentage (%) |
| --- | --- | --- |
| **Age groups in years (N=435)** |  |  |
| 15 – 19 | 40 | 9.2 |
| 20 – 24 | 116 | 26.7 |
| 25 – 29 | 108 | 24.8 |
| 30 – 34 | 88 | 20.2 |
| 35 – 49 | 83 | 19.1 |
| **Women education level (N=435)** |  |  |
| No education | 83 | 19.1 |
| Primary | 222 | 51.0 |
| Secondary and higher | 130 | 29.9 |
| **Marital status (N=435)** |  |  |
| Not in union | 33 | 7.6 |
| Married | 302 | 69.4 |
| Cohabiting | 100 | 23.0 |
| **Has a marriage certificate in their union (n=302)** |  |  |
| No | 178 | 58.9 |
| Yes | 124 | 41.1 |
| **Duration of union in years (n = 435)** |  |  |
| <5 | 159 | 36.6 |
| 5 – 9 | 128 | 29.4 |
| 10 and above | 148 | 34.0 |
| **Place of Residence (N=435)** |  |  |
| Urban | 120 | 27.6 |
| Rural | 315 | 72.4 |
| **Household wealth index (N=435)** |  |  |
| Poor | 183 | 42.1 |
| Middle | 90 | 20.6 |
| Rich | 162 | 37.2 |

| **Respondent Characteristics** | **Frequency** | **Percentage (%)** |
| --- | --- | --- |
| **Women's working status (N=435)** |  |  |
| Not working or unemployed | 199 | 45.8 |
| Working or employed | 236 | 54.3 |
| **Number of Children (N=435)** |  |  |
| No child | 65 | 14.9 |
| 1 child | 103 | 23.7 |
| 2-3 children | 144 | 33.1 |
| 4 or more children | 123 | 28.3 |
| **Acceptance of wife beating (N=435)** |  |  |
| Do not accept | 397 | 91.3 |
| Accept | 38 | 8.7 |
| **Reported partner's age (n=402)** |  |  |
| 19 – 24 | 39 | 9.7 |
| 25 – 34 | 187 | 46.5 |
| 35 – 95 | 176 | 43.8 |
| **Reported partner education level (n=402)** |  |  |
| No education | 53 | 13.2 |
| Primary | 206 | 51.2 |
| Secondary and higher | 131 | 32.6 |
| Don't know | 12 | 3.0 |
| **Reported partner occupation (n=402)** |  |  |
| Not working | 30 | 7.5 |
| Professionals | 31 | 7.7 |
| Sales, clerks and others | 46 | 11.4 |
| Agriculture | 121 | 30.1 |
| Skilled manual | 84 | 20.9 |
| Unskilled manual | 90 | 22.4 |
| **Reported partners who use alcohol (n=402)** |  |  |
| Do not use alcohol | 321 | 79.9 |
| Use alcohol | 81 | 20.1 |
| **Partner reported to have another wife (n=402)** |  |  |
| Have no other wife | 334 | 83.1 |
| Have other wives | 62 | 15.4 |
| don't know | 6 | 1.5 |

**n\*\*** sample size varies across variables, as some of the respondents were not in union although they were pregnant

### Prevalence of Intimate Partner Violence

The overall prevalence of IPV among pregnant women was 27.46% (95% CI: 22.94–32.50). Emotional violence was the most common form (25.26%), followed by sexual (11.04%) and physical (11.01%) violence. Regional variations were substantial, with IPV prevalence highest in Mara (60.3%), Songwe (50.1%), and Singida (39.0%), while Lindi (0%) and Ruvuma (4%) reported the lowest rates.

**Table 2:** The prevalence of IPV among pregnant women aged 15 - 49 years in Tanzania from the 2022 TDHS-MIS.

| Respondent's Characteristics | Frequency (%) | Frequency with IPV (%) in the sample | Weighted IPV Prevalence % | 95% CI for the weighted prevalence |
| --- | --- | --- | --- | --- |
| **Respondents** | **435 (100)** | 102 (23.4) | 27.4 | 22.94 - 32.50 |
| **Age groups (n=435)** |  |  |  |  |
| 15 – 19 | 40 (9.2) | 8 (20.0) | 32.6 | 15.16 – 56.72 |
| 20 – 24 | 116 (26.7) | 31 (26.7) | 27.7 | 18.94 – 38.60 |
| 25 – 29 | 108 (24.8) | 22 (20.4) | 23.9 | 15.11 – 35.62 |
| 30 – 34 | 88 (20.2) | 22 (25.0) | 30.2 | 19.55 – 43.43 |
| 35 – 49 | 82 (19.1) | 19 (22.9) | 25.5 | 15.32 – 39.35 |
| **Women's education level (n=435)** |  |  |  |  |
| No education | 83 (19.1) | 18 (21.7) | 25.1 | 16.47 - 36.32 |
| Primary | 222 (51.0) | 61 (27.5) | 28.6 | 22.51 - 35.60 |
| Secondary and Higher | 130 (29.0) | 23 (17.7) | 26.5 | 17.75 - 37.66 |
| **Marital status (n=435)** |  |  |  |  |
| Not in union | 33 (7.6) | 9 (27.2) | 33.6 | 16.32 - 56.73 |
| Married | 302 (69.4) | 67 (22.2) | 26.6 | 20.37 - 33.99 |
| Cohabiting | 100 (23.0) | 26 (26.0) | 27.5 | 18.38 - 39.00 |
| **Has a marriage certificate for the current union (n=402)** |  |  |  |  |
| No | 178 (58.9) | 50 (28.1) | 31.7 | 23.6 – 41.09 |
| Yes | 124 (41.1) | 17 (13.7) | 14.3 | 8.04 – 24.1 |
| **Duration of union in years (n=435)** |  |  |  |  |
| <5 | 159 (36.6) | 28 (17.6) | 22.8 | 14.73 – 33.56 |
| 5 – 9 | 128 (29.4) | 35 (27.3) | 33.1 | 23.52 – 44.26 |
| 10 and above | 148 (34.0) | 39 (26.4) | 28.7 | 22.94 – 52.98 |
| **Place of Residence (n=435)** |  |  |  |  |
| Urban | 120 (27.6) | 24 (20.0) | 19.6 | 12.36 - 29.62 |
| Rural | 315 (72.4) | 78 (24.8) | 30.1 | 25.20 - 35.58 |
| **Women's wealth index (n=435)** |  |  |  |  |
| Poor | 183 (42.1) | 53 (29.0) | 32.6 | 25.08 – 41.05 |
| Middle | 90 (20.6) | 21 (23.3) | 25.2 | 15.82 – 37.67 |
| Rich | 162 (37.2) | 28 (17.3) | 22.4 | 14.07 – 33.85 |

| **Respondent's Characteristics** | **Frequency (%)** | **Frequency with IPV (%) in the sample** | **Weighted IPV Prevalence %** | **95% CI for the weighted prevalence** |
| --- | --- | --- | --- | --- |
| **Respondents** | **435 (100)** | 102 (23.4) | 27.4 | 22.94 - 32.50 |
| **Women's working status (n=435)** |  |  |  |  |
| Not working or unemployed | 199 (45.8) | 35 (17.6) | 22.1 | 15.59 - 30.34 |
| Working or employed | 236 (54.3) | 67 (28.4) | 31.7 | 24.82 - 39.59 |
| **Number of children (n=435)** |  |  |  |  |
| No child | 65 (14.9) | 15 (23.1) | 34.4 | 19.70 - 52.79 |
| 1 child | 103 (23.7) | 15 (14.6) | 14.9 | 7.65 - 26.94 |
| 2 -3 children | 144 (33.1) | 42 (29.2) | 35.4 | 25.88 - 46.27 |
| 4 or more children | 123 (28.3) | 30 (24.4) | 25.4 | 17.96 - 34.63 |
| **Acceptance of wife beating (n=435)** |  |  |  |  |
| Do not accept | 397 (91.3) | 85 (21.4) | 25.5 | 20.71 - 30.95 |
| Accept | 38 (8.7) | 17 (50.0) | 45.7 | 29.63 - 62.66 |
| **Reported partners' age groups in years (n=402)** |  |  |  |  |
| 19 – 24 | 39 (9.7) | 8 (20.5) | 25.9 | 12.15 - 46.82 |
| 25 – 34 | 187 (46.5) | 42 (22.5) | 26.5 | 18.99 - 35.61 |
| 35 – 95 | 176 (43.8) | 43 (24.4) | 27.7 | 20.35 - 36.37 |
| **Reported partner's education level (n=402)** |  |  |  |  |
| No formal education | 53 (13.2) | 11 (20.75) | 25.1 | 13.42 - 42.07 |
| Primary | 206 (51.2) | 60 (29.1) | 31.2 | 24.44 - 38.94 |
| Secondary and Higher | 131 (32.6) | 19 (14.5) | 19.2 | 10.12 - 33.47 |
| Don't know | 12 (3.0) | 3 (25.0) | 15.8 | 3.6 – 40.0 |
| **Reported partner's occupation (n=402)** |  |  |  |  |
| Not working | 30 (7.5) | 6 (20.0) | 17.1 | 6.44 - 38.32 |
| Professionals | 31 (7.7) | 6 (19.4) | 24.6 | 6.96 - 58.85 |
| Sales clerks and others | 46 (11.4) | 8 (17.4) | 20.4 | 12.21 - 60.64 |
| Agriculture | 121 (30.1) | 28 (23.1) | 24.1 | 17.11 - 32.87 |
| Skilled manual | 84 (20.9) | 20 (23.8) | 29.2 | 19.30 - 41.67 |
| Unskilled manual | 90 (22.4) | 25 (27.8) | 35.4 | 24.06 - 48.59 |
| **Reported partner's alcohol use status (n=402)** |  |  |  |  |
| No | 321 (79.9) | 62 (17.3) | 22.0 | 17.05 - 27.94 |
| Yes | 81 (20.1) | 40 (49.4) | 50.0 | 37.63 - 62.37 |
| **Reported partner having other wives (n=402)** |  |  |  |  |
| No other wife | 334 (83.1) | 69 (20.7) | 23.8 | 18.80 - 29.67 |
| Have other wives | 62 (15.4) | 23 (37.1) | 45.2 | 27.06 - 60.41 |
| Don't know | 6 (1.5) | 2 (33.3) | 31.8 | 16.24 - 52.80 |

**n\*\*** sample size varies across variables, as some of the respondents were not in union although they were pregnant

### Factors Associated with IPV

In the multivariable model, partner alcohol uses increased IPV risk more than twofold (APR = 2.55; 95% CI: 1.50–4.31). Partner having other wives was associated with higher IPV prevalence (APR = 1.75; 95% CI: 1.11–2.87). Union duration of 5–9 years was significantly associated with higher IPV risk (APR = 2.65; 95% CI: 1.14–6.18). Protective factors included having a marriage certificate (APR = 0.51; 95% CI: 0.28–0.92), but also having at least one child (APR = 0.40; 95% CI: 0.17–0.95).

**Table 3:**
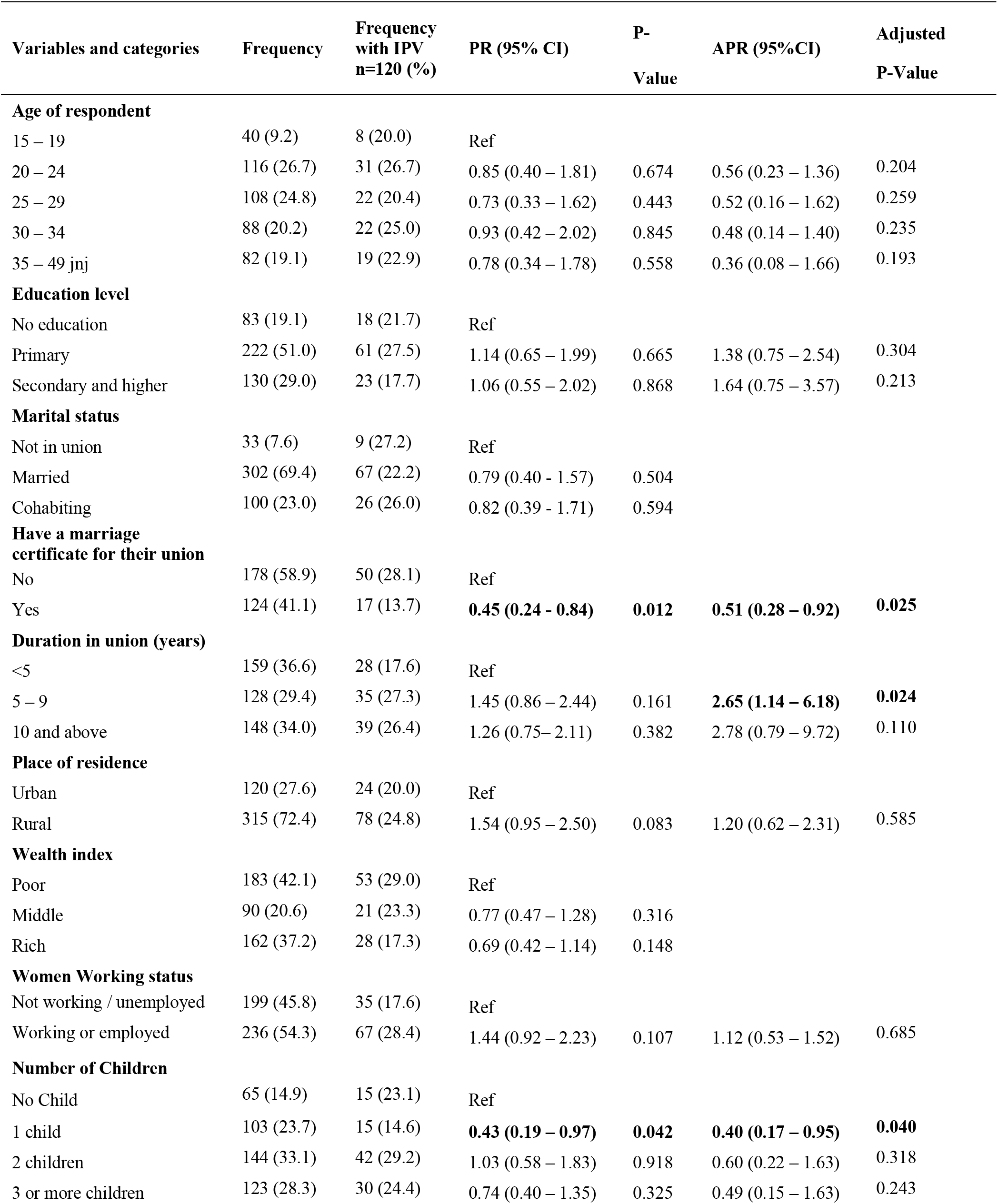
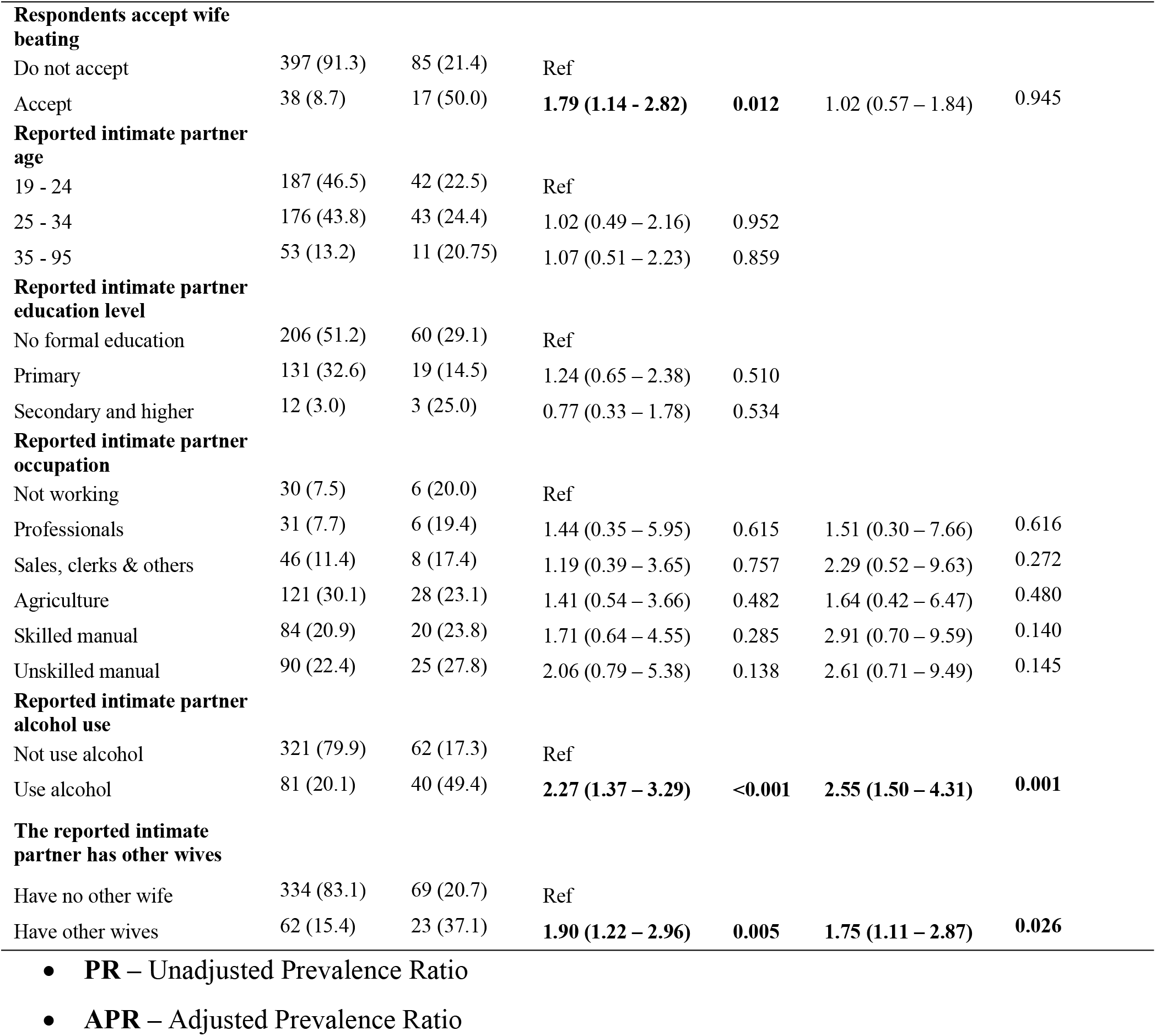
Factors Associated with IPV Among Pregnant Women in Tanzania from the 2022 TDHS-MIS.

| Variables and categories | Frequency | Frequency with IPV<br>n=120 (%) | PR (95% CI) | P-Value | APR (95%CI) | Adjusted P-Value |
| --- | --- | --- | --- | --- | --- | --- |
| **Age of respondent** |  |  |  |  |  |  |
| 15 – 19 | 40 (9.2) | 8 (20.0) | Ref |  |  |  |
| 20 – 24 | 116 (26.7) | 31 (26.7) | 0.85 (0.40 – 1.81) | 0.674 | 0.56 (0.23 – 1.36) | 0.204 |
| 25 – 29 | 108 (24.8) | 22 (20.4) | 0.73 (0.33 – 1.62) | 0.443 | 0.52 (0.16 – 1.62) | 0.259 |
| 30 – 34 | 88 (20.2) | 22 (25.0) | 0.93 (0.42 – 2.02) | 0.845 | 0.48 (0.14 – 1.40) | 0.235 |
| 35 – 49 jnj | 82 (19.1) | 19 (22.9) | 0.78 (0.34 – 1.78) | 0.558 | 0.36 (0.08 – 1.66) | 0.193 |
| **Education level** |  |  |  |  |  |  |
| No education | 83 (19.1) | 18 (21.7) | Ref |  |  |  |
| Primary | 222 (51.0) | 61 (27.5) | 1.14 (0.65 – 1.99) | 0.665 | 1.38 (0.75 – 2.54) | 0.304 |
| Secondary and higher | 130 (29.0) | 23 (17.7) | 1.06 (0.55 – 2.02) | 0.868 | 1.64 (0.75 – 3.57) | 0.213 |
| **Marital status** |  |  |  |  |  |  |
| Not in union | 33 (7.6) | 9 (27.2) | Ref |  |  |  |
| Married | 302 (69.4) | 67 (22.2) | 0.79 (0.40 - 1.57) | 0.504 |  |  |
| Cohabiting | 100 (23.0) | 26 (26.0) | 0.82 (0.39 - 1.71) | 0.594 |  |  |
| **Have a marriage certificate for their union** |  |  |  |  |  |  |
| No | 178 (58.9) | 50 (28.1) | Ref |  |  |  |
| Yes | 124 (41.1) | 17 (13.7) | **0.45 (0.24 - 0.84)** | **0.012** | **0.51 (0.28 – 0.92)** | **0.025** |
| **Duration in union (years)** |  |  |  |  |  |  |
| <5 | 159 (36.6) | 28 (17.6) | Ref |  |  |  |
| 5 – 9 | 128 (29.4) | 35 (27.3) | 1.45 (0.86 – 2.44) | 0.161 | **2.65 (1.14 – 6.18)** | **0.024** |
| 10 and above | 148 (34.0) | 39 (26.4) | 1.26 (0.75– 2.11) | 0.382 | 2.78 (0.79 – 9.72) | 0.110 |
| **Place of residence** |  |  |  |  |  |  |
| Urban | 120 (27.6) | 24 (20.0) | Ref |  |  |  |
| Rural | 315 (72.4) | 78 (24.8) | 1.54 (0.95 – 2.50) | 0.083 | 1.20 (0.62 – 2.31) | 0.585 |
| **Wealth index** |  |  |  |  |  |  |
| Poor | 183 (42.1) | 53 (29.0) | Ref |  |  |  |
| Middle | 90 (20.6) | 21 (23.3) | 0.77 (0.47 – 1.28) | 0.316 |  |  |
| Rich | 162 (37.2) | 28 (17.3) | 0.69 (0.42 – 1.14) | 0.148 |  |  |
| **Women Working status** |  |  |  |  |  |  |
| Not working / unemployed | 199 (45.8) | 35 (17.6) | Ref |  |  |  |
| Working or employed | 236 (54.3) | 67 (28.4) | 1.44 (0.92 – 2.23) | 0.107 | 1.12 (0.53 – 1.52) | 0.685 |
| **Number of Children** |  |  |  |  |  |  |
| No Child | 65 (14.9) | 15 (23.1) | Ref |  |  |  |
| 1 child | 103 (23.7) | 15 (14.6) | **0.43 (0.19 – 0.97)** | **0.042** | **0.40 (0.17 – 0.95)** | **0.040** |
| 2 children | 144 (33.1) | 42 (29.2) | 1.03 (0.58 – 1.83) | 0.918 | 0.60 (0.22 – 1.63) | 0.318 |
| 3 or more children | 123 (28.3) | 30 (24.4) | 0.74 (0.40 – 1.35) | 0.325 | 0.49 (0.15 – 1.63) | 0.243 |

| Variables and categories | Frequency | Frequency with IPV n=120 (%) | PR (95% CI) | P-Value | APR (95%CI) | Adjusted P-Value |
| --- | --- | --- | --- | --- | --- | --- |
| **Respondents accept wife beating** |  |  |  |  |  |  |
| Do not accept | 397 (91.3) | 85 (21.4) | Ref |  |  |  |
| Accept | 38 (8.7) | 17 (50.0) | **1.79 (1.14 - 2.82)** | **0.012** | 1.02 (0.57 – 1.84) | 0.945 |
| **Reported intimate partner age** |  |  |  |  |  |  |
| 19 - 24 | 187 (46.5) | 42 (22.5) | Ref |  |  |  |
| 25 - 34 | 176 (43.8) | 43 (24.4) | 1.02 (0.49 – 2.16) | 0.952 |  |  |
| 35 - 95 | 53 (13.2) | 11 (20.75) | 1.07 (0.51 – 2.23) | 0.859 |  |  |
| **Reported intimate partner education level** |  |  |  |  |  |  |
| No formal education | 206 (51.2) | 60 (29.1) | Ref |  |  |  |
| Primary | 131 (32.6) | 19 (14.5) | 1.24 (0.65 – 2.38) | 0.510 |  |  |
| Secondary and higher | 12 (3.0) | 3 (25.0) | 0.77 (0.33 – 1.78) | 0.534 |  |  |
| **Reported intimate partner occupation** |  |  |  |  |  |  |
| Not working | 30 (7.5) | 6 (20.0) | Ref |  |  |  |
| Professionals | 31 (7.7) | 6 (19.4) | 1.44 (0.35 – 5.95) | 0.615 | 1.51 (0.30 – 7.66) | 0.616 |
| Sales, clerks & others | 46 (11.4) | 8 (17.4) | 1.19 (0.39 – 3.65) | 0.757 | 2.29 (0.52 – 9.63) | 0.272 |
| Agriculture | 121 (30.1) | 28 (23.1) | 1.41 (0.54 – 3.66) | 0.482 | 1.64 (0.42 – 6.47) | 0.480 |
| Skilled manual | 84 (20.9) | 20 (23.8) | 1.71 (0.64 – 4.55) | 0.285 | 2.91 (0.70 – 9.59) | 0.140 |
| Unskilled manual | 90 (22.4) | 25 (27.8) | 2.06 (0.79 – 5.38) | 0.138 | 2.61 (0.71 – 9.49) | 0.145 |
| **Reported intimate partner alcohol use** |  |  |  |  |  |  |
| Not use alcohol | 321 (79.9) | 62 (17.3) | Ref |  |  |  |
| Use alcohol | 81 (20.1) | 40 (49.4) | **2.27 (1.37 – 3.29)** | **<0.001** | **2.55 (1.50 – 4.31)** | **0.001** |
| **The reported intimate partner has other wives** |  |  |  |  |  |  |
| Have no other wife | 334 (83.1) | 69 (20.7) | Ref |  |  |  |
| Have other wives | 62 (15.4) | 23 (37.1) | **1.90 (1.22 – 2.96)** | **0.005** | **1.75 (1.11 – 2.87)** | **0.026** |

- **PR** – Unadjusted Prevalence Ratio - **APR** – Adjusted Prevalence Ratio

## DISCUSSION

Intimate partner violence (IPV) affects nearly one in four pregnant women in Tanzania, with emotional violence being most common. The factors with significant associations included having a marriage certificate, having one child, partner alcohol use, partner who had man other women, longer union duration, and a history of childhood exposure to domestic violence. No significant association was found between IPV and socio-domestic factors such as age, education, occupation, or place of residence.

The overall IPV prevalence in this study is lower than estimates from several regional analyses. Regional disparities within Tanzania were notable, with the highest IPV prevalence in Mara, Songwe, and Singida, and the lowest in Ruvuma, Shinyanga, and Kigoma, while no case was reported in Lindi. Such differences may reflect variations in cultural norms, gender relations, and reporting behaviors across regions. The overall prevalence in this study was lower than that reported by the 2022 TDHS -MIS (39%) for all women aged 15 to 49 years(1). while an analysis of Ethiopian Demographic and Health Survey (DHS) data reported a prevalence of 28.74% (2). Considering a multilevel analysis of sub-Saharan Africa, the prevalence of IPV among pregnant women was 41.94% (3) which is higher than that from our findings. The study findings reported the prevalence was higher among adolescent similar to that done in Tanzania among young women where it was found high prevalence of IPV among adolescents(4) Similarly, a study on the magnitude and determinants of IPV in East Africa reported the prevalence of 32.66% among pregnant women(5).

The lower IPV prevalence among pregnant women found in this study may be attributed to cultural and legal differences, improved gender-based violence policies in Tanzania, variations in survey methods, underreporting, and differing access to support services. But also it seems that women at health facilities were more able to disclose their violence status than during the population-based survey, as observed in other studies from Northern Tanzania found the prevalence of 30.3% among the pregnant women attending antenatal care in Moshi health facilities(6).

Emotional violence was the most commonly reported form of intimate partner violence among pregnant women in our study followed by physical and sexual IPV which reported at nearly equal rates, but with a lower rate. This pattern mirrors findings from the 2022 TDHS-MIS report, where emotional IPV was reported at 27% and sexual IPV at 11%(1). Comparable trends have been reported across sub-Saharan Africa, where emotional violence remains the predominant form of IPV during pregnancy(3,7).

These findings highlight the considerable burden of emotional IPV during pregnancy and emphasize the need for IPV prevention and screening strategies that prioritize emotional violence, which is often under-recognized yet has significant mental and physical health consequences for women and their unborn children(8). It is often more frequent, socially tolerated, and normalized in many communities, making it more likely to be reported. Pregnant women also feel more comfortable disclosing emotional IPV as it carries less stigma and a lower risk of retaliation than physical or sexual violence. Additionally, emotional violence may increase during pregnancy due to increased stress, hormonal changes, partner jealousy, or financial pressure, while some partners may reduce physical or sexual violence out of concern for the fetus.

Several relationship and partner-level characteristics emerged as strong predictors of IPV. Partner alcohol consumption was one of the most significant factors, increasing IPV risk more than two folds. This findings aligns with evidence from Kenya, Rwanda and Ethiopia, where alcohol use by male partners is consistently linked to higher IPV risk(7,9–11).

Parity and IPV association was observed also in a study done in East Africa, where it was discussed that, presence of one child may necessitate ongoing partner contact and act as protection against IPV, and if the number of children increases, the expenditure of the household also increases, therefore acting as factor for IPV(12). Alcohol consumption may heighten aggression, impair judgement, and exacerbate household conflict. Addressing alcohol misuse through community-based interventions, male engagement, and counseling programs could reduce IPV risk during pregnancy.

Similarly, women whose partners had other wives were nearly twice the prevalence to experience IPV compared to those in monogamous during pregnancy. Polygamous marriages often intensify competition, jealous and economic strain, leading to conflict and diminished women’s bargaining power. These findings are consistent with others studies in Tanzania and Kenya, which reported that polygamy is a major driver of IPV due to patriarchal power dynamics and unequal resource distribution (4,10). Interventions promoting gender equity and responsible partnership could help mitigates IPV risk in such unions.

The duration of the marital union also influenced IPV prevalence. Women who had been in unions for 5 to 9 years were more than twice as likely to report IPV compared to those in shorter unions. This suggests that IPV may increase over time, as conflicts accumulate or as violence becomes normalized within relationships. Similar trends have been reported in Ethiopia, where longer union duration was associated with increased IPV experience during pregnancy by their intimate partners(13). Continuous relationship counseling and early interventions may therefore be essential to prevent escalation of abuse in long-term unions.

## STUDY STRENGTHS AND LIMITATIONS

This study used nationally representative data and a robust analytical approach, enhancing the generalizability of its findings. However, the reliance on self-reported data introduces information bias that may influence reliability of the study findings primarily because of underreporting due to fear, stigma, or recall bias. The cross-sectional design also limits causal inference. Despite these limitations, the findings provide valuable evidence to guide policy and intervention strategies aimed at reducing IPV during pregnancy in Tanzania.

## CONCLUSION

This study found that almost 1 in 4 pregnant women in Tanzania experience intimate partner violence (IPV), with emotional IPV being the most common form. Key protective factors included having a marriage certificate, and having one child, while major factors associated with IPV among pregnant women were partner alcohol use, a partner who had married other wives, and longer union duration. No significant association was found between IPV and socio-demographic factors such as age, education, occupation, or place of residence.

## RECOMMENDATIONS

1. Ministry of Health, regional and council health management teams to strengthen IPV screening and support services in maternal healthcare by integrating it in routine antenatal care (ANC) services across all regions, especially in Mara, Songwe, and Singida.
2. The government of Tanzania should strengthen legal and social support for formal marriage by collaborating with local government and community organizations to increase access to marriage registration, particularly in rural areas. Public awareness campaigns should highlight the legal protections afforded by marriage certificates, such as the right to property and resources against abuse.
3. The Ministry of Health, in collaboration with the Ministry of Community Development, Gender, Women, and Special Groups, including developing partners that are involving in the prevention and control of violence against women, should strengthen a behavior change program for men who are involved in alcoholism and polygamous marriages to prevent the cycle of violence.
4. Community leaders and local government authorities should promote social norms change and support networks by addressing polygamy and promote family stability through community sensitization and marital counseling.
5. Civil society organization should focus on women’s empowerment, education and advocacy programs that improve awareness of rights, financial independence and decision-making

## Data Availability

The data underlying the results presented in the study are available from: https://www.dhsprogram.com/data/dataset_admin/login_main.cfm.

## ACKNOWLEDGEMENT

Above all, I am sincerely grateful to Almighty God for His continual guidance, strength, and blessings. I extend my heartfelt appreciation to the Department of Epidemiology and Biostatistics at MUHAS for supervision and mentorship, the Centers for Disease Control and Prevention (CDC) under the Tanzania Field Epidemiology and Laboratory Training Program, the Directorate of Reproductive and Child Health at the Ministry of Health for their support, and the Demographic and Health Survey (DHS) Program for their contribution in this study.

## LIST OF ABBREVIATIONS

ANC: Antenatal Care
APR: Adjusted Prevalence Ratio
CI: Confidence Interval
DHS: Demographic and Health Survey
EAs: Enumeration Areas
FELTP: Field Epidemiology and Laboratory Training Program
GBD: Global Burden of Disease
GBV: Gender-Based Violence
IPV: Intimate Partner Violence
MUHAS: Muhimbili University of Health and Allied Sciences
NIMR: National Institute of Medical Research
SDG: Sustainable Development Goals
TDHS-MIS: Tanzania Demographic and Health Survey and Malaria Indicator Survey
WHO: World Health Organization

## Authors contributions

Conceptualization and design: Cephlen Mathayo, Rose Mpembeni, Rogath S. Kishimba, Habib Ismail

Data extraction; Cephlen Mathayo, Jasmine Chilembu,

Analysis and interpretation; Cephlen Mathayo, Agnes Tesha, Rose Mpembeni, Rogath S. Kishimba, Habib Ismail, Godson Ngowi

Drafting and review of manuscript; Cephlen Mathayo, Rose Mpembeni and Rogath S. Kishimba, Agnes Tesha

All authors revised the manuscript and approved the final version.

